# Validation of semi-automatic citation screening software for creating clinical practice guidelines: A protocol for a prospective observational study

**DOI:** 10.1101/2022.11.17.22282374

**Authors:** Takehiko Oami, Yohei Okada, Tatsuma Fukuda, Masaaki Sakuraya, Taka-aki Nakada, Nobuaki Shime

**Affiliations:** Department of Emergency and Critical Care Medicine, Chiba University Graduate School of Medicine, Chiba, Japan; Department of Preventive Services, Kyoto University Graduate School of Medicine, Kyoto, Japan; Health Services and Systems Research, Duke-NUS Medical school, National University of Singapore, Singapore; Department of Emergency and Critical Care Medicine, Toranomon Hospital, Tokyo, Japan; Department of Emergency and Intensive Care Medicine, JA Hiroshima General Hospital, Hatsukaichi, Japan; Department of Emergency and Critical Care Medicine, Graduate School of Biomedical and Health Sciences, Hiroshima University, Hiroshima, Japan

## Abstract

**Background:** This study aims to investigate the quality of the literature search and workload saving using the semi-automatic software for citation screening in the development of the Japanese Clinical Practice Guidelines for Management of Sepsis and Septic Shock (J-SSCG).

**Methods:** We will conduct a prospective study to compare the efficiency of citation screening between the conventional method using Rayyan and semi-automatic citation screening using ASReview. The two independent reviewers will conduct literature searches for clinical questions. During the session, we objectively measure the time to accomplish the citation screening. After the citation screening, we will calculate the sensitivity and specificity from the results of the conventional and semi-automatic procedures. Also, we will compare the accumulated time between the two methods.

**Trial registration:** This research is submitted with the University hospital medical information network clinical trial registry (UMIN-CTR) [UMIN000049366].

**Conflicts of interest:** All authors declare no conflicts of interest to have.

**Funding:** None

## Background

A systematic review of the literature is essential in updating or collecting the latest evidence among the several components of developing clinical practice guidelines [1]. However, the process needs an iterative screening of a vast number of publications, which is often a time-consuming and intensive workload for reviewers [2, 3]. Recently, several automation software for citation screening using machine learning has been developed [4-8]. Based on the training data loaded into the software by researchers, the software is expected to predict the relevant publications with reliability and high accuracy, which could result in workload savings for citation screening in the process of systematic review [9].

Despite the potential advantages, such machine learning-based automatic screening has not been applied to the systematic review in the development of guidelines. Barriers to the application of automatic screening potentially include the lack of trust, difficulty in setting, inappropriate selection of the software, incompatibility with conventional methods, and the variation of usability in different search fields [9, 10]. Those concerns could be resolved through meticulous investigations and effective application; however, the feasibility and validity of semi-automatic screening software using machine learning have been rarely investigated in the development of clinical practice guidelines. Therefore, it is warranted to evaluate the usefulness of the software for citation screening in the guidelines.

We hypothesized that the semi-automatic tools of citation screening would demonstrate the sufficient quality of the literature search and reduce the workload of researchers in the development of Japanese Clinical Practice Guidelines for Management of Sepsis and Septic Shock (J-SSCG). In the process of conducting citation screening, we will aim to compare the accuracy and workload of semi-automated software with those of manual literature screening.

## Methods

### Study design and settings

We will conduct a prospective study for the validation of semi-automatic screening software following steps: selection of the citation screening tools which could be applicable with the committee members, setting of the clinical questions, and independent performance of the citation screening to evaluate the semi-automatic citation screening tool compared to the conventional method in the process of developing J-SSCG 2024. We submitted the review protocol to the pre-print server (medRxiv) and registered with the University hospital medical information network (UMIN) clinical trial registry [UMIN UMIN000049366].

### Clinical questions in the J-SSCG

Previously, J-SSCG 2020 was published by the Japanese Society of Intensive Care Medicine (JSICM) and the Japanese Association for Acute Medicine (JAAM), considering Japanese-specific clinical settings for sepsis and septic shock [11]. The revised version of J-SSCG 2020 will be created in 2024 as J-SSCG 2024.

We will evaluate the efficiency of the automated software using the clinical questions (CQs) that were newly selected in the guidelines. Comprehensive literature reviews will be conducted using CENTRAL, PubMed, and Ichushi-Web for the CQs. The working group members will define the search strategy, confirming that no key studies are missing. Only Japanese and English literature will be selected. After downloading all the titles and abstracts, members will integrate the files from different search engines and remove duplicated files in the citation manager. We will use EndNote (Clarivate Analytics, USA) as citation manager software in J-SSCG 2024. The five CQs (Table 1) were optimal to investigate as targets because the timing of the conventional citation screening fit the progress of this study.

**Table 1.**
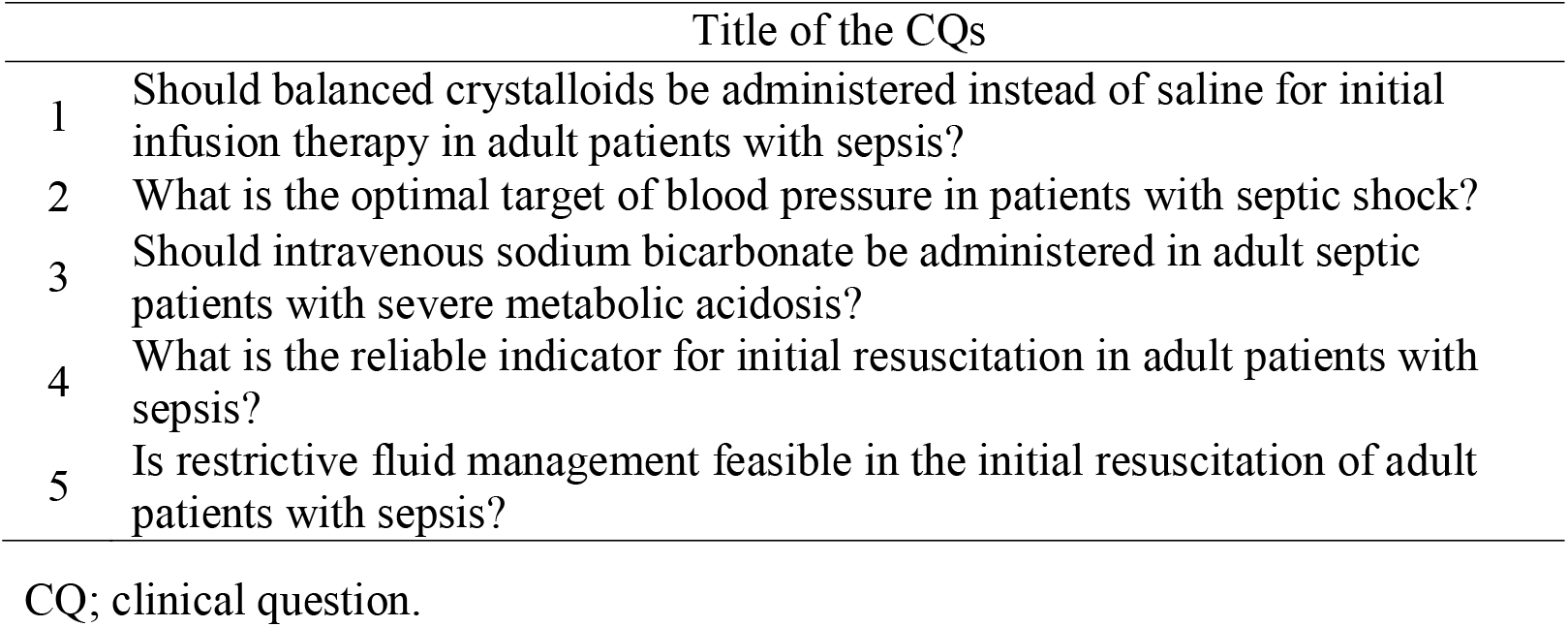
The list of enrolled clinical questions.

### Automatic citation screening

We will use ASReview (https://doi.org/10.1038/s42256-020-00287-7) as semi-automatic software for citation screening to verify the efficiency and feasibility in guidelines. To select automated software for the citation screening, we screened application software that had been published in the past according to the following conditions: 1) open source software with high reproducibility and applicability 2) software development using machine learning. The usability and compatibility with other software for systematic reviews were evaluated based on user-friendliness for working group members in J-SSCG (Fig. S1).

ASReview is semi-automated software for citation screening using machine learning-based text-mining technology. Before the procedure, reviewers will need to import the dataset from citation managers. We will use the same dataset with the procedure of the conventional tool for citation screening. The process of exporting references is the same as the conventional method for citation screening. The software requires at least one relevant study and non-relevant records labeled by reviewers to predict the relevance of the remaining publications using the training data. In this study, we will provide all the key studies as relevant literature and label 100 randomly selected records. According to the relevance of the references, the software will rank the records and export the file containing the results. Two independent reviewers (T.O. and Y.O.) will conduct a semi-automated citation screening using the software. at the same time. Each reviewer will indicate the threshold among the ranked records to determine the relevant references. After the session, the two reviewers will confirm there is no conflict between the results.

### Conventional citation screening

The processed files in EndNote will be exported to Rayyan (https://rayyan.qcri.org/welcome), the systematic review assistant software. Two independent reviewers will judge the publication as a relevant or an irrelevant study based on the title and abstract. They will discuss the judgement to resolve the conflict or the third person will conduct a review in case of the conflict. After the first screening using the title and abstract, the second screening will be conducted using full-text articles. We will only cover the first screening for the research subject in this study.

### Data collection

We will collect the following information about reviewers: age, sex, professional category, education degree, specialty, duration of clinical experiences, total number of systematic reviews. We will compare the accuracy and workload between the conventional literature screening and the semi-automation of citation screening software. In this study, we will only assess the feasibility of the software using the first screening session. To evaluate the accuracy, we will count the number of missing or redundant references between the conventional citation screening and semi-automatic procedure. As an assessment of the workload, we will record the duration of the session for the citation screening. For ASReview, reviewers will record the video during the session to conduct citation screening. Then, the third person (T.N.) will measure the total time from the beginning of the review to the end of the session to secure the objectivity. Since Rayyan has a function to measure the time of the literature screening, we will record the time from the beginning of the screening to the end of the session. At the time of this study protocol, systematic reviews for the new CQs have not been conducted yet.

### Statistical analysis

The continuous variables are means (standard deviation) or medians (interquartile ranges [IQR]), as appropriate. Using the number of references extracted from the manual and automated software systematic reviews, we will calculate the sensitivity and specificity of the results of conventional citation screening as the gold standard. The cumulative time will be calculated from the session of systematic review in each CQ. GraphPad Prism 9 (GraphPad Software, San Diego, CA, USA) will be used for the statistical analysis.

## Data Availability

All data produced in the present study are available upon reasonable request to the authors.

## Figure legends

**Supplemental Figure S1:**
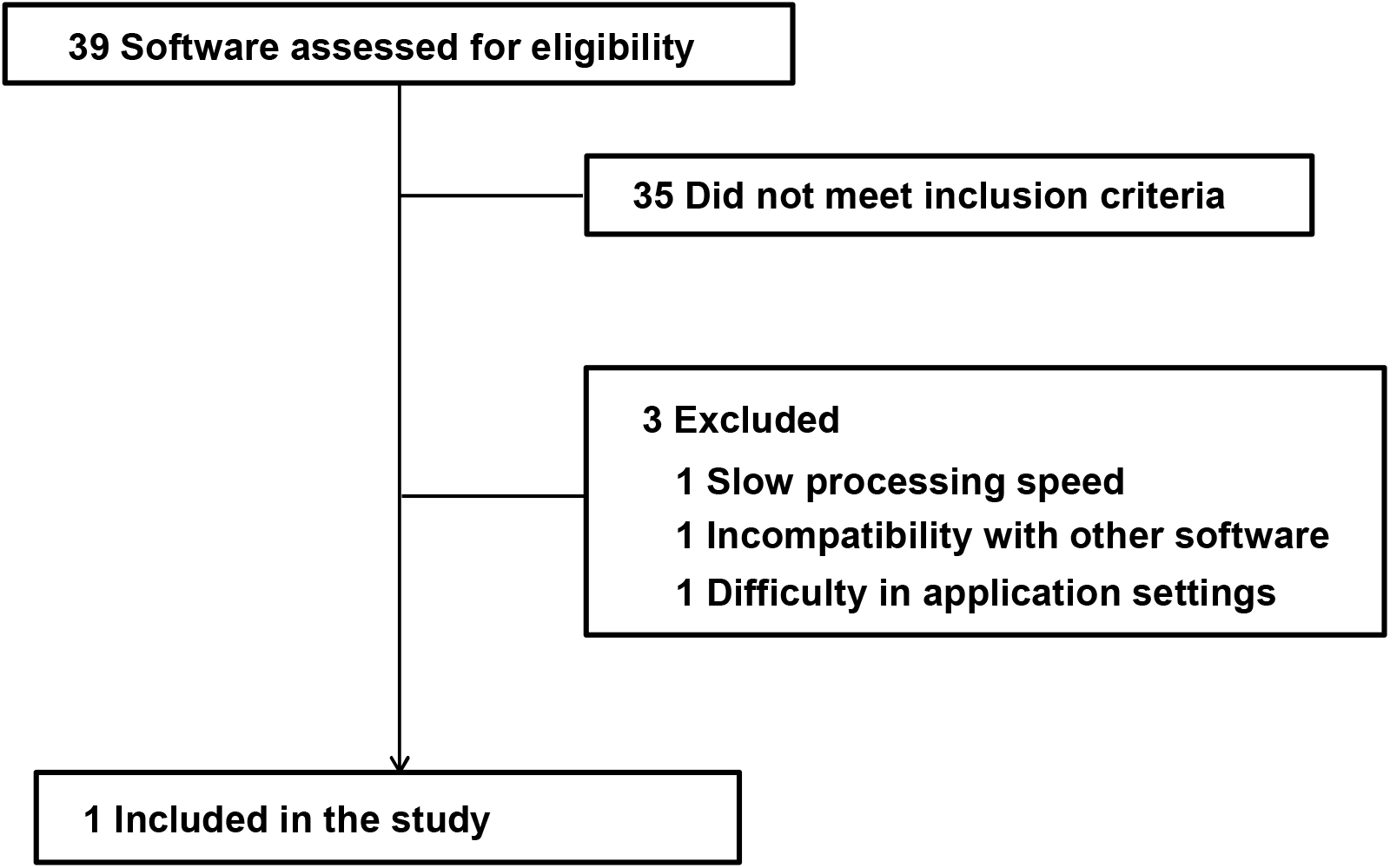
Flowchart of the software selection in this study.

