## Supplementary figures and images for "Validation of semi-automatic citation screening software for creating clinical practice guidelines: A protocol for a prospective observational study"

### Supplemental Figure S1

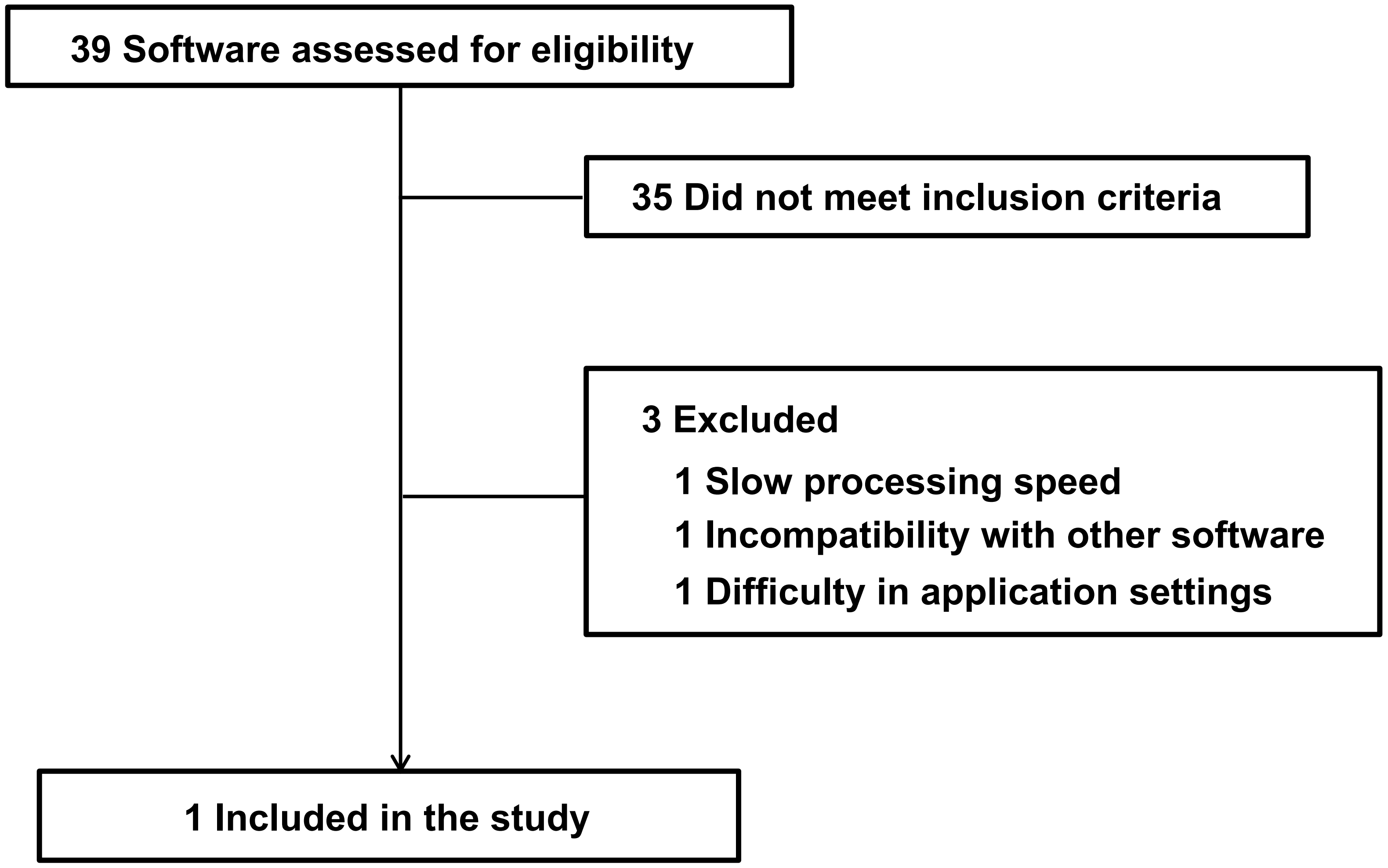
